# PanelAppRex aggregates disease gene panels and facilitates sophisticated search

**DOI:** 10.1101/2025.03.20.25324319

**Authors:** Quant Group, Simon Boutry, Ali Saadat, Sinisa Savic, Luregn J. Schlapbach, Jacques Fellay, Dylan Lawless

## Abstract

**Motivation:** Gene panel data are essential for variant interpretation and genomic diagnostics, but existing resources are fragmented, inconsistently annotated, and not easily accessible for programmatic use. We developed PanelAppRex, a harmonised dataset and interactive search tool that integrates over 58,000 curated gene-disease panel associations. It supports natural language-style queries by gene, phenotype, disease group, and mode of inheritance (MOI), with results returned in machine-readable export formats.

**Results:** The resulting dataset includes standardised gene identifiers, disease annotations, MOI, and literature support, enabling seamless integration into bioinformatic pipelines. We benchmarked fifteen case studies spanning immunology, neurology, and additional disease areas. Under the recommended usage, in which the union of returned panels is considered, the causal gene was recovered in every case. Across all returned panels, the causal gene was present in 85.6% of panels. For manual interface interpretation, the causal gene was present in the user-selected best-fit panel(s) in all fifteen benchmarked cases.

**Availability:** The platform data is openly available at PanelAppRex base [Data set], Zenodo https://doi.org/10.5281/zenodo.15736689, with source code at https://github.com/DylanLawless/PanelAppRex, and demonstration page at https://panelapprex.github.io/landing_page. The dataset is maintained for a minimum of two years following publication.

## 1 Introduction

Disease-gene panels are widely used in both clinical and research settings to support the diagnosis and interpretation of genetic disorders. These panels provide structured lists of genes known to be associated with specific phenotypes or disease groups, helping clinicians prioritise candidate variants during genomic analysis. Sources like Genomics England (GE)’s PanelApp and PanelApp Australia host comprehensive, expert-curated panels that are actively maintained to reflect new genetic knowledge (1). For instance, these resources are integral to the NHS National Genomic Test Directory and the 100,000 Genomes Project (1). Despite their utility, these panel datasets are distributed across multiple platforms and formats, and are difficult to aggregate programmatically without data loss or inconsistency. Manual selection, interpretation, and cross-referencing of gene panels remain labour intensive, especially when integrating with other variant-level annotations from genomic resources.

Accurate causal variant interpretation requires consistent integration of structured annotations from key resources such as GE’s PanelApp, ClinVar, UniProt, and Ensembl (1–4). To address this, we developed PanelAppRex, a panel aggregation and machine-readable dataset with core gene identifiers required for gene-panel linking, together with structured annotations including inheritance modes, disease terms, and supporting evidence where available. The dataset is suitable for integration into AI-based workflows and downstream variant interpretation pipelines. In parallel, PanelAppRex provides a natural language-style search interface that streamlines discovery by supporting queries based on gene names, phenotypes, disease groups, and other key attributes. We further demonstrate how the structured core resource can be extended with an AI-derived summary layer for interpretation and downstream Retrieval-Augmented Generation (RAG)-style workflows.

## 2 Methods

### 2.1 Data

The PanelAppRex core model contained 58,592 entries containing annotation fields, including the gene name, disease-gene panel ID, disease-related features, confidence measurements. Data from gnomAD v4 comprised 807,162 individuals, including 730,947 exomes and 76,215 genomes (5). This dataset provided 786,500,648 single nucleotide variants and 122,583,462 indels, with variant type counts of 9,643,254 synonymous, 16,412,219 missense, 726,924 nonsense, 1,186,588 frameshift and 542,514 canonical splice site variants. ClinVar data were obtained from the variant summary dataset available from the NCBI FTP site, and included 6,845,091 entries, which were processed into 91,319 gene classification groups and a total of 38,983 gene classifications (2). Data from Ensembl was sourced for validation of identifiers such as gene IDs and Human Genome Organisation Gene Nomenclature Committee (HGNC) symbols (4). Disease interactions were compared against GE’s PanelApp (1).

### 2.2 Implementation

PanelAppRex was implemented in R and integrated data from the sources listed above. Reference data was merged for comparison with the core model in two formats: a simplified version (Panel ID, Gene) and a complex version including metadata such as confidence level, Mode of Inheritance (MOI), and disease information, and several metadata summary statistics. In addition, the tool incorporated a search module to execute complex user queries. The search functionality supports queries by gene names, phenotypes, disease names, disease groups, panel names, genomic locations and other identifiers.

### 2.3 Usage

We provide both a pre-compiled user interface and an analysis-ready dataset. On a desktop browser, the user interface allows for queries via the integrated search bar in HyperText Markup Language (HTML), where a JavaScript function splits the query into individual terms and progressively filters all entries, retaining only those that match all active terms while ignoring unmatched ones. This enables users to perform complex, partial matching queries (e.g. “paediatric *RAG1* primary immunodeficiency skin disorder”) to rapidly identify the panel most closely associated with their hypothesis on primary immunodeficiency and paediatric skin disorders. After querying and selecting a panel, the complete underlying data is returned.

The analysis-ready dataset contains the dataset in multiple file formats. Our recommended strategy is a bioinformatic approach which can use the union of returned panels, or to apply a scoring system to rank panels in multi-group analyses. Bioinformatically, users can import the provided ready-for-use datasets in Tab-Separated Values (TSV) or R Data Serialization format (RDS) formats. A typical use case might involve users merging with their own omic data based on gene or Ensembl ID. The following code snippet, available in minimal_example.R, demonstrates how to load the data in R:

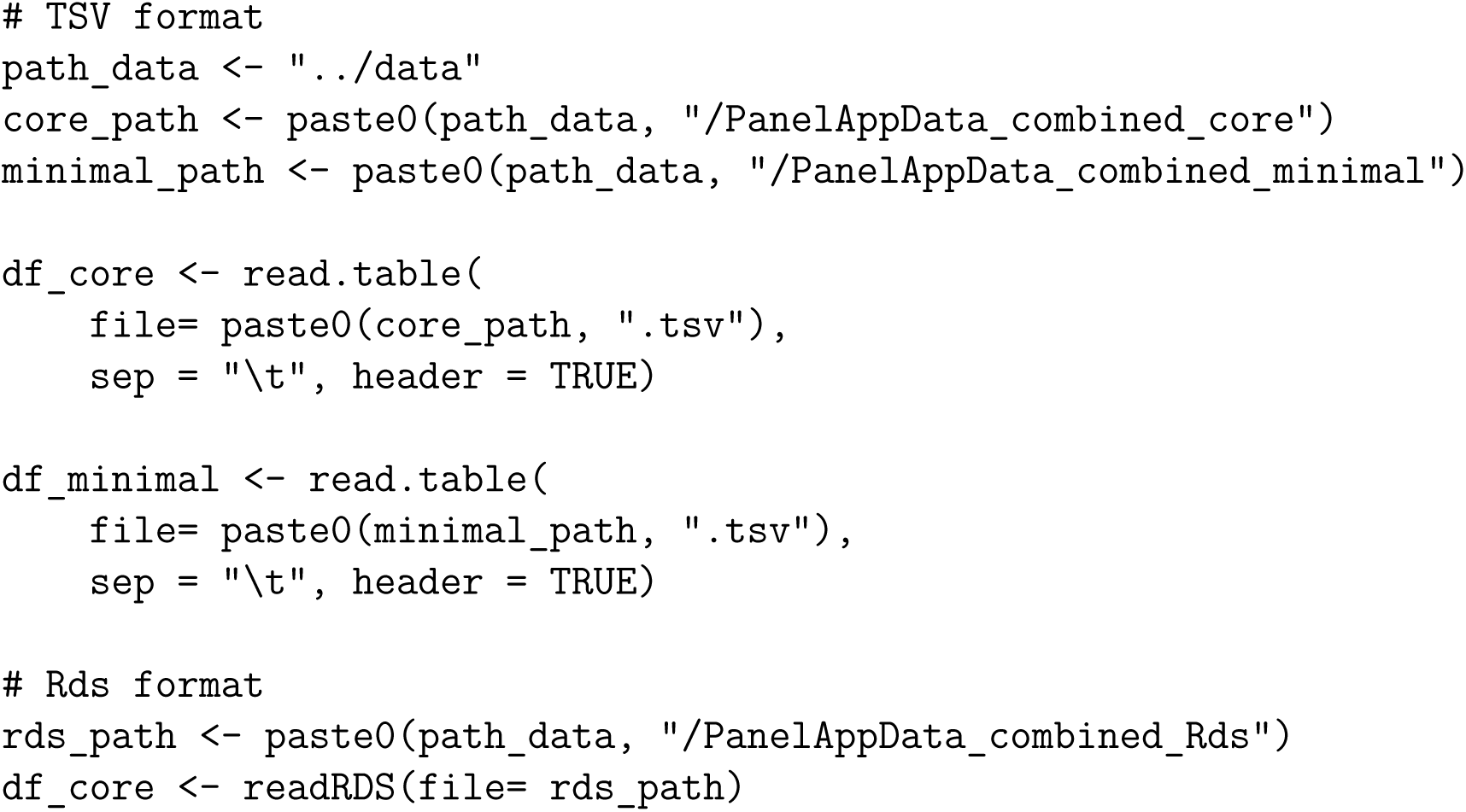

### 2.4 Validation for completeness in core data

To ensure the reliability and completeness of the core dataset, we systematically assessed whether key gene-level fields were present for each entry after merging the core dataset. Specifically, we checked for non-missing values in gene symbols, associated publications, disease panel names, MOI, and Online Mendelian Inheritance in Man (OMIM) gene IDs. Where HGNC or Ensembl gene IDs were missing, we used programmatic queries to the Ensembl database via the ‘biomaRt’ R package to recover the identifiers using the available HGNC symbol as input (6). This validation step was applied to the full integrated PanelAppRex dataset.

### 2.5 Benchmarking for manual queries

To mimic a clinician attempting to diagnose a genetic disease, we systematically selected published genetic case reports. We chose the first five eligible case reports from each of three sources: Journal of Allergy and Clinical Immunology (JACI), the journal Neurology, and a pre-defined cross-disciplinary PubMed search, yielding a total of fifteen case studies across phenotypes.

For the cross-disciplinary set, PubMed was searched using the query “(genetic) AND (case report[Title])”, restricted to the last twelve months and Free Full Text (accessed February 2026). We then screened the results for clinical genetic case reports using exome or genome analysis and selected the first five eligible studies. The combined set allowed us to assess the method across immunological, neurological, and additional disease areas in a small benchmark set.

For each case report, we used only the clinical narrative presented before any genetic analysis or molecular diagnosis was introduced in the original publication. Using the same procedure across studies, phenotype terms were extracted from this pre-genetic clinical description and ranked from most to least specific or relevant. Query construction was completed before checking the PanelAppRex results. No causal gene names were added unless they appeared explicitly in the original clinical description, and no additional synonyms or aliases were introduced manually.

For the unusually long fifth case study, which described five individual patients, we used an OpenAI o3-mini model only to standardise extraction of phenotype keywords from the pre-genetic clinical case description text. No information from the genetic results or discussion was provided to the model, and the extracted terms were then ranked using the same procedure as for the other case studies.

The ranked phenotype terms were then used as keyword queries, simulating a naïve clinical starting point. Search terms were added incrementally until no panels matched. We then assessed whether PanelAppRex retrieved at least one panel containing the reported causal gene for each case study. For manual interface use, we also recorded the user-selected best-fit panel(s). This was a judgement-based assessment intended to reflect plausible user choice, defined as the returned panel or panels considered most clinically aligned with the pre-genetic case description. Where several returned panels appeared similarly appropriate, more than one panel could be recorded. The full list of included studies, source links, input queries, returned panels, user-selected best-fit panel choices, and benchmark outputs is provided in **Table S1**. Benchmarking analyses were performed on the core dataset; the AI-derived demonstration “Info” field described below was not used in this evaluation.

### 2.6 Applications in AI-based RAG

To demonstrate how the core PanelAppRex dataset can be expanded for AI-assisted interpretation, we generated an optional panel-level summary layer suitable for down-stream RAG-style workflows. For each gene, we retrieved mechanistic text from the UniProtKB human proteome and merged these summaries with PanelAppRex panel membership and metadata, including panel name, disease group, MOI, and reported phenotypes, to create a context dataset for each panel. We then applied a GPT-based model (gpt-4.1-mini, OpenAI; temperature 0.1) with a fixed prompt requesting a concise clinician-facing description of shared mechanisms and phenotypes across the panel gene set. Across all processed panels, this involved approximately 7 million input tokens and 2 million output tokens. The model produced three structured outputs per panel, consisting of a detailed analysis section, an overview paragraph, and 3 to 6 bullet points. These outputs were stored together with panel identifiers and exposed in the demonstration interface as an experimental panel “Info” field. This AI-derived summary layer was not used for the panel-ranking benchmarks reported in this study.

## 3 Results

### 3.1 Core dataset

PanelAppRex successfully aggregated data for 451 panels to offer a database and user-friendly search functionality (**Figure 1**). Users can retrieve results filtered by gene names, phenotypes, disease groups and other criteria. The browser-based user interface system also provides a table view with panel details and provides options for exporting results in comma-separated values (CSV), Excel, or Portable Document Format (PDF) format. Bioinformatic uses may include generating virtual panels, constructing prior odds, or supporting formal reporting in qualifying variant protocols. Summary statistics are reported in **Figure S2**. Panel reuse varied across genes, with some like *RAG1* appearing in up to 7 distinct panels (**Figure S2 B**). The majority of panels had fewer than 1000 genes. For example, the panel related to Primary Immunodeficiency (PID), also referred to as Inborn Errors of Immunity (IEI), contains 572 genes, while the panel linked to IEI and unexplained death in infancy includes 1675 genes (**Figure S2 C**).

**Figure 1:**
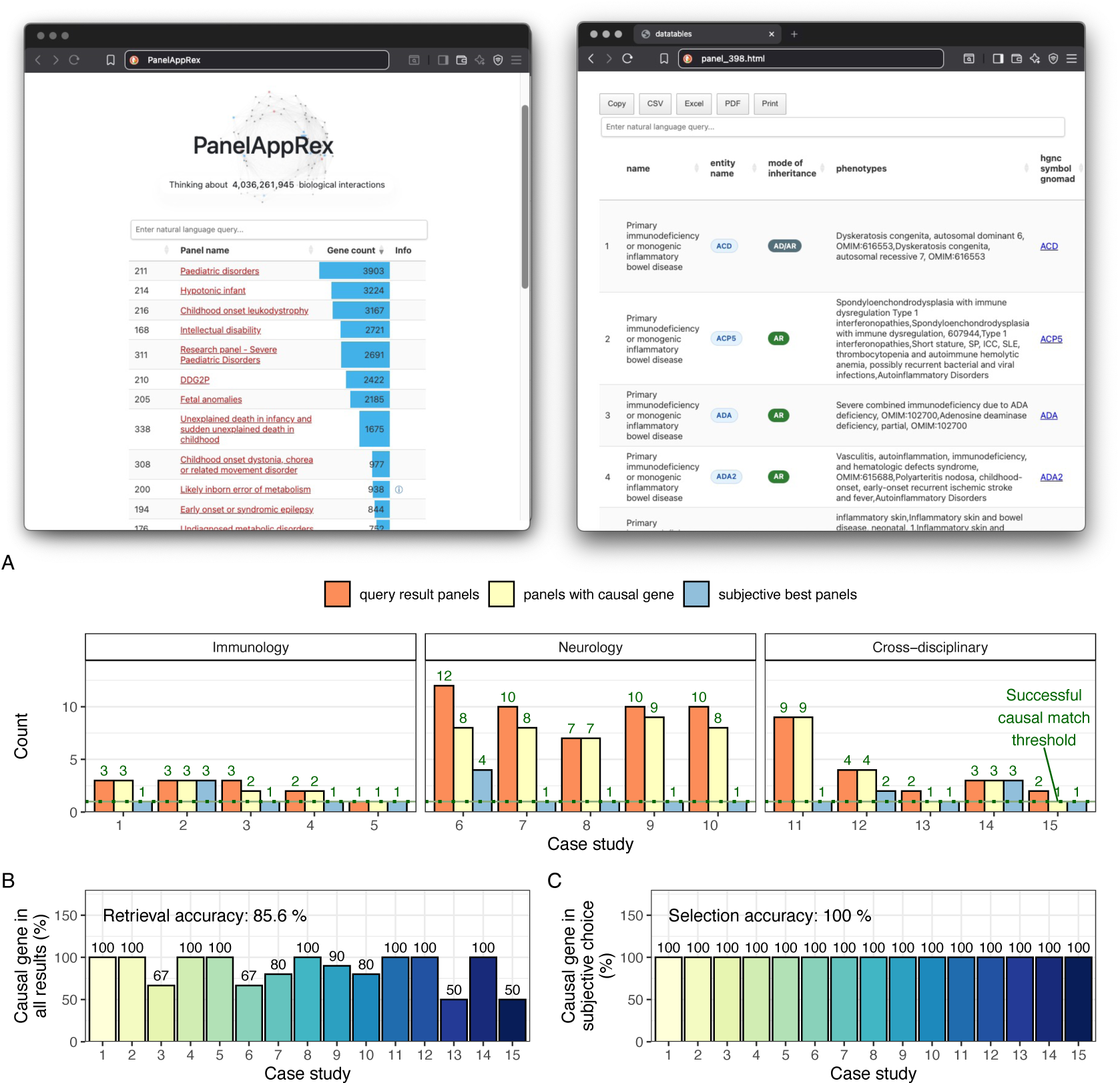
PanelAppRex interface displaying search results for a complex query. Top: screenshots showing the search interface, with the left panel displaying the full database before filtering and the right panel showing detailed results for a selected panel. Bottom: benchmark metrics are presented as follows: (A) for each of the fifteen case studies, the total number of panels returned after query input, the number of panels that included the causal gene, and the user-selected best-fit panel(s); (B) the percentage of all returned panels that included the true causal gene; (C) the percentage of user-selected best-fit panel(s) that contained the true causal gene.

### 3.2 Validation of completeness in the core dataset

To assess whether the dataset was analysis-ready, we audited completeness of key annotation fields required for joining, filtering, and interpretation (**Figure S1**). We report entry-level completeness across all gene-disease panel entries. For HGNC and Ensembl gene identifiers, we additionally used automated recovery of missing IDs from Ensembl because these identifier mappings are not subjective. After recovery, the core identifiers required for gene-panel linking were complete for all valid gene entries. A small number of panel entries do not have gene-specific fields (i.e. Ensembl gene ID) because they are not genes. For example, HPE1 is a phenotype identifier rather than a gene symbol and therefore is not represented in HGNC or Ensembl gene records. Some secondary annotation fields remained incomplete, including MOI, and we did not attempt to fill these in the core dataset because doing so would require non-primary-source decisions better reserved for downstream implementations. The validated dataset therefore provides a stable and analysis-ready identifier layer for downstream integration while preserving the original availability of secondary annotations.

### 3.3 Benchmarking for manual queries confirms accurate retrieval of causal disease genes

To evaluate the practical utility of our system on a desktop browser, we applied the benchmarking approach to fifteen published genetic diagnosis case studies that used exome or genome sequencing. These comprised five immunology-focused PID case reports, five neurology-focused case reports, and five randomly selected domains, as described in Methods. The query process retrieved gene panels using natural language-style input. For example, in the first case study on Hereditary Angioedema, the authors reported suspecting that the condition was linked to “*SERPING1* Factor XII and edema” which we used as the query terms. Although the true causal gene term *F12* (using its official HGNC name) was not mentioned, the inclusion of the alias name and related disease terms enabled the system to retrieve the panel containing *F12* through indexed metadata.

While an expert might readily recognise “Factor XII” as an alias name for *F12*, this correct query result demonstrates that the system can make such connections automatically, based on its extensive metadata, supporting users with varying levels of genetic expertise. Most of the case report queries relied purely on phenotypic descriptions without any candidate genes.

Across the fifteen benchmarked case studies, the system retrieved at least one panel containing the reported causal gene, despite these genes not appearing explicitly in our submitted query terms (**Table S1**).

Under the recommended usage, in which the union of all returned panels is considered, the causal gene was recovered in all fifteen benchmarked cases. Across all returned panels, the causal gene was present in 85.6% of panels. For manual interface interpretation, the causal gene was present in the user-selected best-fit panel(s) in all fifteen benchmarked cases (**Figure 1**).

These metrics were derived by comparing the causal gene identified from each case study to the panels returned by the query. For automated use, we recommend considering the union of returned panels. For manual interface use, we also recorded a user-selected best-fit panel assessment intended to reflect plausible user choice, acknowledging that certain panels, such as the established PID gene panel, may be more clinically aligned than broader, less specific panels (**Table S1**).

Overall, these results suggest that the approach can identify relevant panels in this benchmark setting and may be useful for supporting panel selection in complex diagnostic scenarios.

### 3.4 Applications in AI-based RAG

Across all panels, we used the core PanelAppRex dataset combined with RAG on the UniProtKB human proteome to generate AI-derived panel summaries that captured panel-specific mechanisms and phenotypes conditioned on disease context and gene composition. These structured summaries are exposed in the interface as an experimental panel “Info” field, intended as a demonstration of how the core dataset can be extended for AI-assisted interpretation. In aggregate, this step consumed approximately 7 million input tokens and produced 2 million output tokens while condensing the 6.6 million-word gene knowledge base into approximately 135,000 words of panel-level summaries, corresponding to roughly fifty-fold compression. This summary layer demonstrates how the structured PanelAppRex core can be extended for AI-assisted interpretation and future downstream RAG-style applications. The benchmarking analyses reported here were performed on the core structured dataset and did not depend on this additional AI-derived field.

### 3.5 Structured inheritance annotations reveal overlapping gene-MOI relationships

To assess the distribution of inheritance annotations we summarised all unique gene-MOI combinations across 58,592 entries as displayed in **Figure S3**. Among 6,280 distinct genes, a total of 9,237 unique gene-MOI pairs were identified, reflecting instances where a gene appears with multiple inheritance modes across different panels. The most common annotations were Autosomal recessive (AR) and Autosomal dominant (AD), consistent with their prevalence in monogenic disease panels. These structured inheritance annotations underpin key applications enabled by PanelAppRex, including MOI-filtered search and model-based prior estimation.

## 4 Discussion

PanelAppRex provides an accessible platform for querying, aggregating, and exporting curated disease-gene panel data, with initial benchmarking in published case studies. Designed for both clinical and research use, it simplifies study planning and variant interpretation through a user-friendly interface, while also offering a harmonised dataset for programmatic workflows. The core model integrates over 58,000 panel-gene associations, with complete coverage of the core identifiers required for linking valid gene entries after recovery, alongside structured disease annotations, HGNC symbols, MOI, OMIM-linked identifiers, and supporting publications where available.

Beyond immediate usability, PanelAppRex provides a structured resource that may support future statistical modelling of disease-associated variation. With annotations for gene-disease relationships, the dataset may be useful for future work on estimating prior probabilities of observing variant classifications (e.g. benign, pathogenic) under different disease MOI. This relates to a broader challenge in clinical genetics: the absence of principled priors for variant interpretation that incorporate not only known pathogenic variants (true positives), but also unobserved pathogenic variants (false negatives) and the absence of pathogenic findings (true negatives) (7; 8). By integrating high-resolution allele frequencies from gnomAD (5), curated classifications from ClinVar (2), and structured gene-disease associations (1), PanelAppRex may support future downstream development of quantitative models of genetic risk.

These capabilities extend panel lookup by supporting structured retrieval, filtering, and downstream analysis in diagnostic and research workflows. Probabilistic modelling, uncertainty estimation, and related approaches remain potential down-stream applications.

The dataset may also provide a useful substrate for future AI-assisted approaches to variant interpretation, including probabilistic inference and annotation workflows (9; 10). We included an AI-derived panel-summary layer as an example extension of the structured core resource. It provides concise panel-level context for interpretation and illustrates how PanelAppRex could support future RAG-style and other AI-assisted workflows.

Several limitations should be acknowledged. Not all known coding genes are currently linked to a disease panel; we prioritised high-confidence, traceable annotations over broad but less reliable coverage. Some genes are over-represented across multiple panels due to historical research biases, leading to non-uniform panel enrichment. We benchmarked the approach in both relatively simple gene-phenotype relationships (e.g. PID) and more complex settings such as neurogenetic disorders, but the benchmark remained limited to 15 published case studies. The user-selected best-fit panel assessment was included to reflect plausible manual interface use and should be interpreted as a judgement-based measure rather than a formal objective ranking metric. Broader real-world evaluation will be needed to determine practical benefit across disease areas. Additionally, not all panels returned by queries may be equally informative. For example, during benchmarking, the COVID-19 research panel frequently appeared alongside the PID panel due to overlapping genes. While technically accurate, such panels may be less relevant to users focused on clinical diagnosis of immunodeficiency. In bioinformatic workflows, these choices can be refined systematically or excluded using filters or downstream logic. In case study three, for example, a disease-based query returned three panels, one of which did not contain the causal gene. This is expected, as broader classifications may include non-specific panels, hindering sensitivity. However, the query was still considered successful in this benchmark framework, because the causal gene appeared in the combined result set, consistent with the recommended bioinformatic strategy of using the union of returned panels. We note that a pre-compiled offline database is useful in secure computing environments, but it does not provide the same update frequency as well-maintained primary-source APIs. For rapidly changing resources, direct API access can offer more current data than an offline snapshot.

PanelAppRex provides a structured and practical resource for interactive search and programmatic access to curated disease-gene panel information. Future work will focus on expanding supported queries, integrating additional variant types and annotations, and evaluating more advanced applications in variant interpretation and risk estimation.

## Supporting information

Table S1 supplemental benchmarks

## Acronyms

AD: Autosomal dominant
AR: Autosomal recessive
API: Application Programming Interface
CSV: comma-separated values
GE: Genomics England
HGNC: Human Genome Organisation Gene Nomenclature Committee
IEI: Inborn Errors of Immunity
IEM: Inborn Errors of Metabolism
HTML: HyperText Markup Language
JACI: Journal of Allergy and Clinical Immunology
MOI: Mode of Inheritance
OMIM: Online Mendelian Inheritance in Man
PDF: Portable Document Format
PID: Primary Immunodeficiency
RAG: Retrieval-Augmented Generation
RDS: R Data Serialization format
TSV: Tab-Separated Values

## Acknowledgements

ClinVar asks its users who distribute or copy data to provide attribution to them as a data source in publications and websites (2). The use of data from UniProt was based on Creative Commons Attribution 4.0 International (CC BY 4.0). We acknowledge GE for providing publicly accessible PanelApp data. The use of data from GE PanelApp was for research purposes and is not for healthcare or commercial services. The use of GE PanelApp source code was under the Apache License 2.0. The experimental (RAG) resource includes machine-assisted annotations generated using the OpenAI API according to their terms of use. We do not access OMIM data but instead direct readers using gene identifiers linked to OMIM.org record pages for further reading and acknowledge this resource. Ensembl data and code are available without restriction and software is provided under the Apache License 2.0 (4). Ensembl and its satellite sites conform to the EBI terms of use; as such we acknowledge attribution to EMBL-EBI for the use of its data resources and tools.

## Contributions

DL designed and performed analyses and wrote the manuscript. SB, AS, SS, JF, LJS designed analysis and wrote the manuscript. The Quant Group is a collaboration across multiple institutions where authors contribute equally; the members of this project were DL, SB, and AS.

## Ethics statement

This study only used data which was previously published and publicly available, as cited in the manuscript. This SwissPedHealth study, under which this work was carried out, was approved based on the advice of the ethical committee Northwest and Central Switzerland (EKNZ, AO_2022-00018). The study was conducted in accordance with the Declaration of Helsinki.

## Data availability

- Data: Lawless, D. (2025). PanelAppRex base Zenodo. https://doi.org/10. 5281/zenodo.15736689
- Source code: https://github.com/DylanLawless/PanelAppRex
- Demonstration page: https://panelapprex.github.io/landing_page.

## 5 Supplemental

Table S1: **Summary of case study queries and PanelAppRex results.** We recommend automated use that accounts for the union of all query results. For manual interface use, judgement-based panel selection is naturally expected. We therefore recorded the user-selected best-fit panel(s), defined as the returned panel or panels judged most clinically aligned with the pre-genetic case description and most likely to be prioritised by a user. In this context, broader but less specific panels (for example, “COVID-19 research” or “Paediatric disorders”) may be deprioritised relative to panels whose title and scope more closely match the presenting condition. The results are summarised in **Figure 1**.

**Figure S1:**
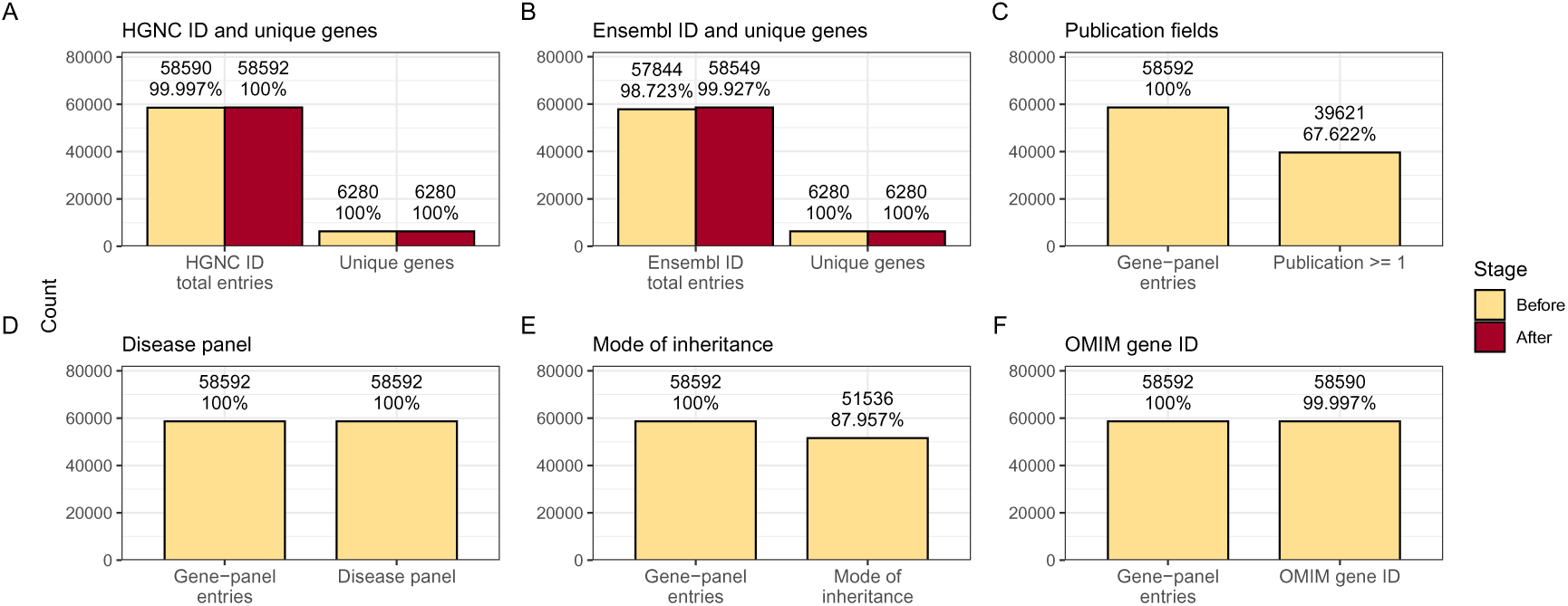
Validation of completeness for major annotation fields in the core dataset. (A) Entry-level completeness of HGNC identifiers before and after recovery using Ensembl biomaRt with HGNC symbol input, shown together with the total number of unique genes in the dataset as a reference. (B) Entry-level completeness of Ensembl gene identifiers before and after recovery using Ensembl biomaRt with HGNC symbol input, shown together with the total number of unique genes in the dataset as a reference. (C) Entry-level completeness of publication annotations, shown as the number of gene-panel entries compared to those with at least one linked publication. (D) Entry-level completeness of disease panel annotations, shown as the number of gene-panel entries with a gene and the number with a disease panel name. (E) Entry-level completeness of mode of inheritance annotations, shown as the number of gene-panel entries with a gene and the number with a recorded mode of inheritance. (F) Entry-level completeness of OMIM gene identifier annotations, shown as the number of gene-panel entries with a gene and the number with an OMIM gene identifier. For all panels the percentages are calculated relative to total gene-panel entries, except for panels A and B where the additional “unique genes” bars are calculated relative to the total number of unique genes. Entry-level refers to the initial merging of data from independent sources.

**Figure S2:**
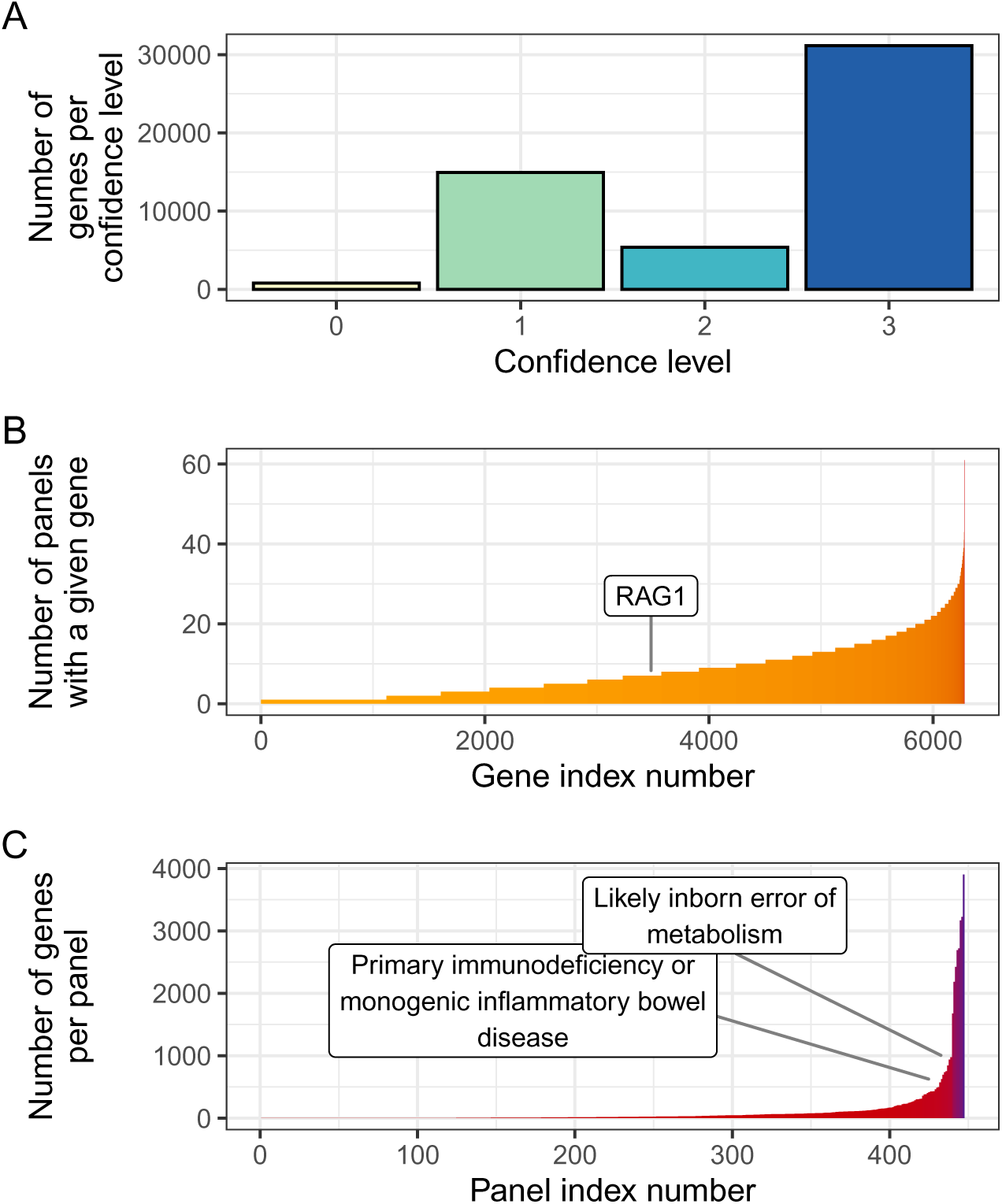
Summary of gene confidence levels, reuse across panels, and panel sizes in the PanelAppRex dataset. (A) Number of genes per confidence level as reported by **GE!**. (B) Number of panels in which each gene is included, with the example gene *RAG1* highlighted to demonstrate that it is present in 7 panels. (C) Number of genes per panel, with two representative panels annotated: PID/IEI and IEM.

**Figure S3:**
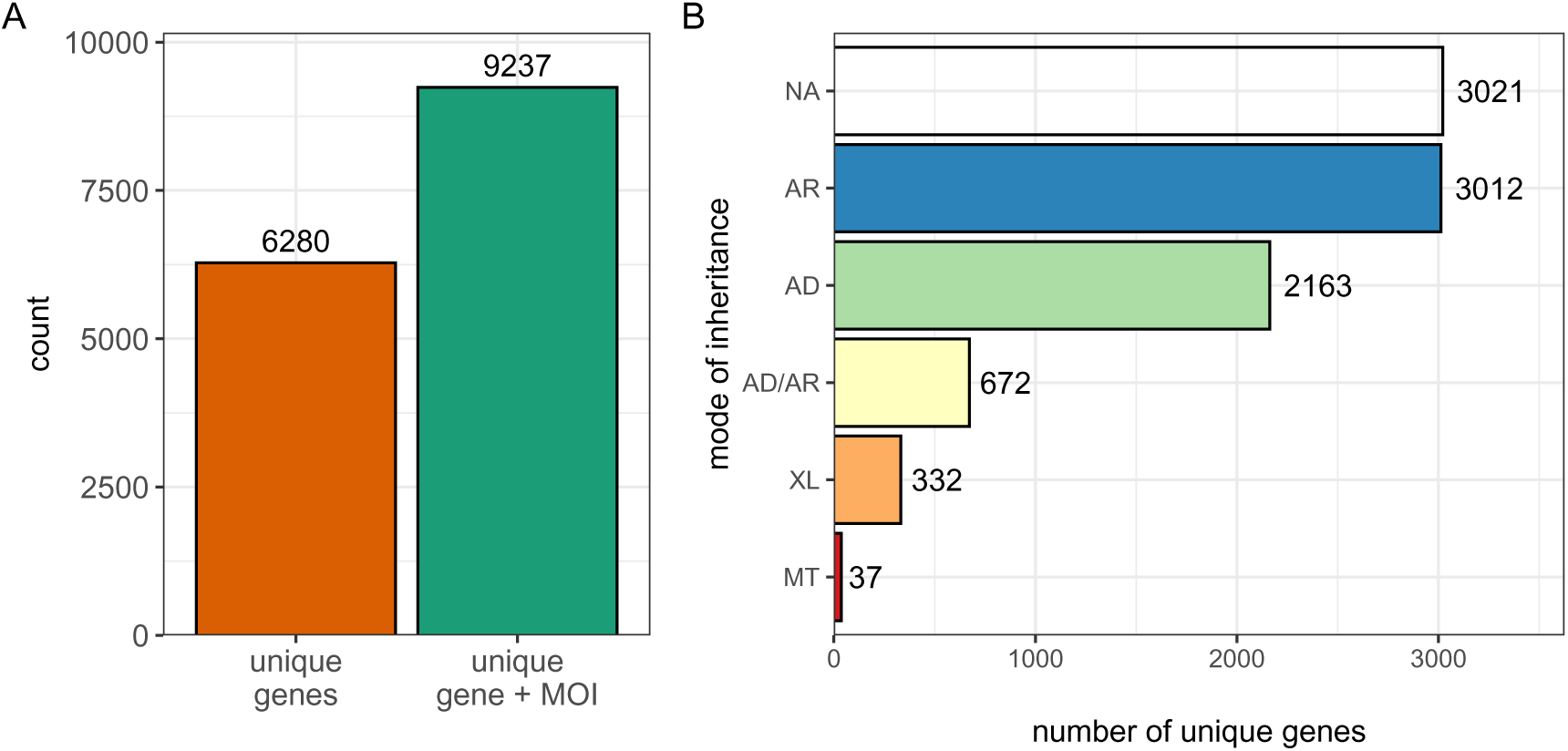
Summary of mode of inheritance annotations in the PanelAppRex dataset. (A) Counts of unique genes annotated with each MOI, based on non-redundant gene-MOI combinations. (B) Total number of unique genes and total number of gene-MOI combinations in the harmonised PanelAppRex dataset.

### 5.1 Performance

To assess whether PanelAppRex improves practical query performance beyond direct use of a comparable resource, we performed a cold-start benchmark against a best-case GE PanelApp Application Programming Interface (API) workflow by re-using the first validation case query, “*SERPING1*, Factor XII, and edema” (**Figure S4**). We used the same query in both settings. This comparison is not fully like-for-like, because the GE PanelApp API does not accept the same phenotype-style query and return a ranked panel result directly. Instead, panel data must first be retrieved and assembled before ranking can begin. To make the comparison as favourable as possible to the API workflow, we deliberately included the known relevant panel from validation case study 1 and then increased the number of queried panels. In practice, a user would not usually know that panel in advance.

PanelAppRex local was faster at every tested subset size for time to final ranked result. The API workflow also required increasing numbers of panel-level requests in order to assemble the returned data before ranking could begin. Queries with more than 100 panel calls resulted in the GE API returning HTTP 429 rate-limiting fair usage errors, so larger panel sets could not be evaluated in this benchmark without introducing additional delays. In addition, the API comparator returned only panel content, whereas PanelAppRex provided the pre-integrated local dataset together with the annotations used elsewhere in the platform such as gene function and disease details.

**Figure S4:**
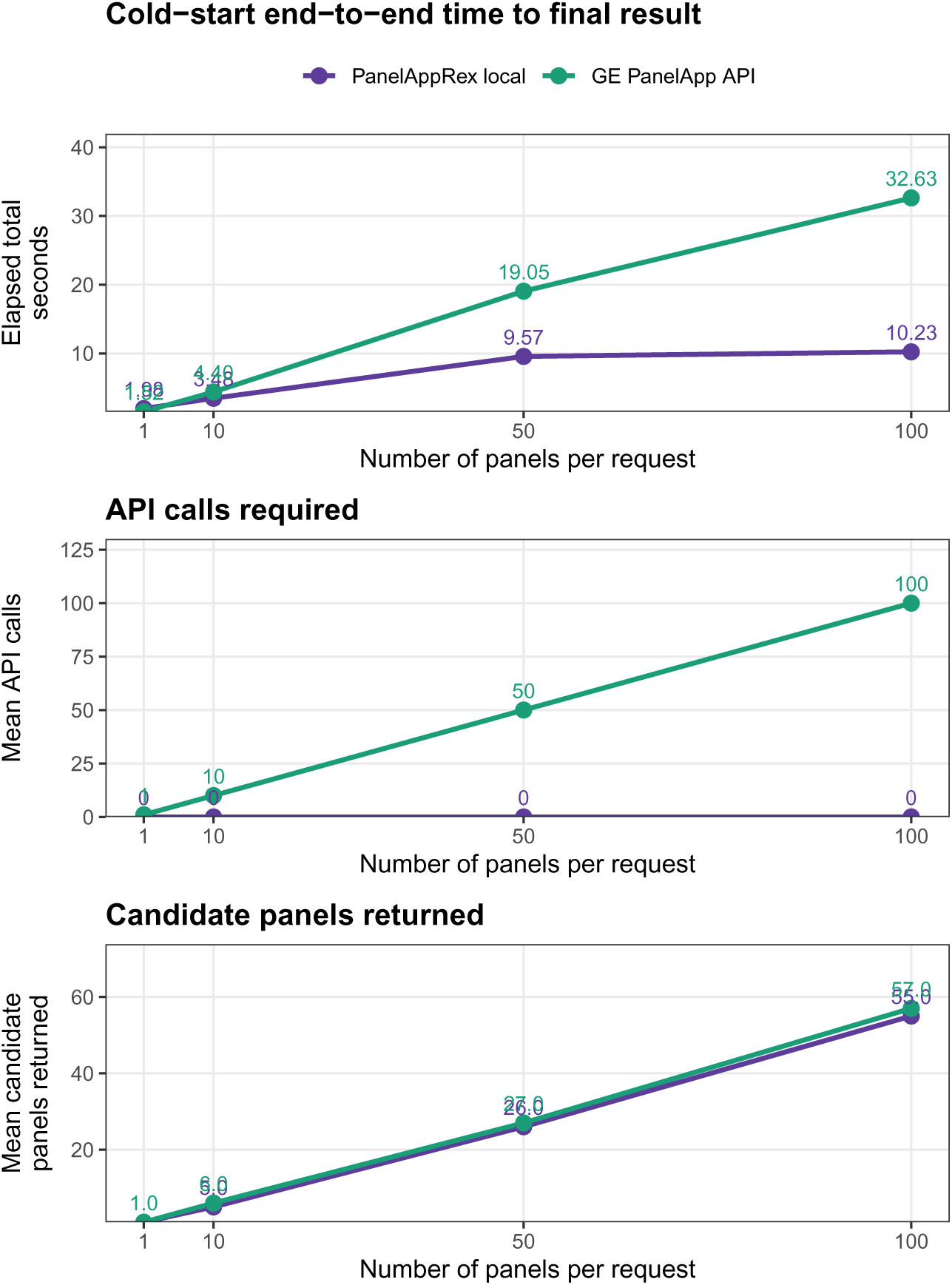
Cold-start benchmark of PanelAppRex local against a best-case GE PanelApp API workflow. The same clinical query from case study 1 was used in both settings. The x axis shows the number of panels in each benchmark set. For each run, the same growing panel set was used in both arms. The API comparator was deliberately made favourable by including the known relevant panel and then increasing the number of additional panels retrieved. (A) End-to-end elapsed time from query to final ranked result. (B) Mean number of API calls required to retrieve and assemble the queried GE panels before ranking. PanelAppRex local required no live API calls at query time. (C) Mean number of candidate panels returned after applying the input query to the retrieved panel set. For this speed benchmark, candidate panels were counted using at least one matching query term. After 150 panels in the benchmark set, the GE API returned HTTP 429 rate-limiting errors, so larger runs could not be completed in this benchmark.

### 5.2 Example AI-based RAG summary

We used the core PanelAppRex dataset combined with RAG on the UniProtKB human proteome to generate AI-derived panel summaries that captured panel-specific mechanisms and phenotypes conditioned on disease context and gene composition. To demonstrate the results here, we show the first panel from the user interface, “Thoracic aortic aneurysm or dissection”, which contains 65 genes. The AI-based RAG “Info” summary for this panel was:

#### Overview

This gene set primarily encompasses disorders of connective tissue and vascular smooth muscle leading to thoracic aortic aneurysm and dissection. Clinical features include progressive dilation and rupture risk of the thoracic aorta, arterial tortuosity, joint hypermobility, skin laxity, skeletal deformities such as scoliosis and pectus excavatum, ocular abnormalities like lens dislocation, and systemic manifestations of connective tissue fragility. Many syndromes represented show autosomal dominant inheritance with variable expressivity and overlap with Marfan, Loeys-Dietz, and Ehlers-Danlos syndromes.

#### Summary

- Defects in collagen synthesis, processing, and fibril assembly compromise connective tissue strength, leading to vascular fragility and skeletal abnormalities.
- Impaired elastic fiber formation and maintenance disrupt aortic wall elasticity, predisposing to aneurysm and dissection.
- Dysregulated TGF-beta signaling pathways alter vascular smooth muscle cell function and extracellular matrix remodeling, contributing to arterial tortuosity and aneurysm formation.
- Smooth muscle contractile apparatus dysfunction affects vascular tone and structural integrity, increasing risk of thoracic aortic aneurysm.
- Phenotypes include thoracic aortic aneurysm and dissection, joint hypermobility, skin hyperextensibility, skeletal deformities (scoliosis, pectus excavatum), and ocular features such as ectopia lentis.
- Syndromic presentations overlap with connective tissue disorders including Loeys-Dietz syndrome, Ehlers-Danlos syndrome (vascular and classic types), and Mar-fan syndrome.

## References

[1] Antonio Rueda Martin, Eleanor Williams, Rebecca E. Foulger, Sarah Leigh, Louise C. Daugherty, Olivia Niblock, Ivone U. S. Leong, Katherine R. Smith, Oleg Gerasimenko, Eik Haraldsdottir, Ellen Thomas, Richard H. Scott, Emma Baple, Arianna Tucci, Helen Brittain, Anna De Burca, Kristina Ibañez, Dalia Kasperaviciute, Damian Smedley, Mark Caulfield, Augusto Rendon, and Ellen M. McDonagh. PanelApp crowdsources expert knowledge to establish consensus diagnostic gene panels. Nature Genetics, 51(11):1560–1565, November 2019. ISSN 1061-4036, 1546-1718. doi: 10.1038/s41588-019-0528-2. URL https://www.nature.com/articles/s41588-019-0528-2.

[2] Melissa J Landrum, Jennifer M Lee, Mark Benson, Garth R Brown, Chen Chao, Shanmuga Chitipiralla, Baoshan Gu, Jennifer Hart, Douglas Hoffman, Wonhee Jang, Karen Karapetyan, Kenneth Katz, Chunlei Liu, Zenith Maddipatla, Adri-ana Malheiro, Kurt McDaniel, Michael Ovetsky, George Riley, George Zhou, J Bradley Holmes, Brandi L Kattman, and Donna R Maglott. ClinVar: improving access to variant interpretations and supporting evidence. Nucleic Acids Research, 46(D1):D1062–D1067, January 2018. ISSN 0305-1048, 1362-4962. doi: 10.1093/nar/gkx1153. URL http://academic.oup.com/nar/article/46/D1/D1062/4641904.

[3] The UniProt Consortium, Alex Bateman, Maria-Jesus Martin, Sandra Orchard, Michele Magrane, Aduragbemi Adesina, Shadab Ahmad, Bowler-Barnett, and others. UniProt: the Universal Protein Knowledgebase in 2025. Nucleic Acids Research, 53(D1):D609–D617, January 2025. ISSN 0305-1048, 1362-4962. doi: 10.1093/nar/gkae1010. URL https://academic.oup.com/nar/article/53/D1/D609/7902999.

[4] Sarah C Dyer, Olanrewaju Austine-Orimoloye, Andrey G Azov, Matthieu Barba, If Barnes, Vianey Paola Barrera-Enriquez, Arne Becker, Ruth Bennett, Martin Beracochea, and others. Ensembl 2025. Nucleic Acids Research, 53(D1):D948–D957, January 2025. ISSN 0305-1048, 1362-4962. doi: 10.1093/nar/gkae1071. URL https://academic.oup.com/nar/article/53/D1/D948/7916352.

[5] Konrad J Karczewski, Laurent C Francioli, Grace Tiao, Beryl B Cummings, Jessica Alföldi, Qingbo Wang, Ryan L Collins, Kristen M Laricchia, Andrea Ganna, Daniel P Birnbaum, et al. The mutational constraint spectrum quantified from variation in 141,456 humans. Nature, 581(7809):434–443, 2020.

[6] Wolfgang Huber Steffen Durinck. biomaRt, 2017. URL https://bioconductor.org/packages/biomaRt.

[7] William B. Hannah, Mitchell L. Drumm, Keith Nykamp, Tiziano Pramparo, Robert D. Steiner, and Steven J. Schrodi. Using genomic databases to determine the frequency and population-based heterogeneity of autosomal recessive conditions. Genetics in Medicine Open, 2:101881, 2024. ISSN 29497744. doi: 10.1016/j.gimo.2024.101881. URL https://linkinghub.elsevier.com/retrieve/pii/S2949774424010276.

[8] Johannes Zschocke, Peter H. Byers, and Andrew O. M. Wilkie. Mendelian inheritance revisited: dominance and recessiveness in medical genetics. Nature Reviews Genetics, 24(7):442–463, July 2023. ISSN 1471-0056, 1471-0064. doi: 10.1038/s41576-023-00574-0. URL https://www.nature.com/articles/s41576-023-00574-0.

[9] John Jumper, Richard Evans, Alexander Pritzel, Tim Green, Michael Figurnov, Olaf Ronneberger, Kathryn Tunyasuvunakool, Russ Bates, Augustin Žídek, Anna Potapenko, Alex Bridgland, Clemens Meyer, Simon A. A. Kohl, Andrew J. Ballard, Andrew Cowie, Bernardino Romera-Paredes, Stanislav Nikolov, Rishub Jain, Jonas Adler, Trevor Back, Stig Petersen, David Reiman, Ellen Clancy, Michal Zielinski, Martin Steinegger, Michalina Pacholska, Tamas Berghammer, Sebastian Bodenstein, David Silver, Oriol Vinyals, Andrew W. Senior, Koray Kavukcuoglu, Pushmeet Kohli, and Demis Hassabis. Highly accurate protein structure prediction with AlphaFold. Nature, 596(7873):583–589, August 2021. ISSN 0028-0836, 1476-4687. doi: 10.1038/s41586-021-03819-2. URL https://www.nature.com/articles/s41586-021-03819-2.

[10] Jun Cheng, Guido Novati, Joshua Pan, Clare Bycroft, Akvilė Žemgulytė, Taylor Applebaum, Alexander Pritzel, Lai Hong Wong, Michal Zielinski, Tobias Sargeant, Rosalia G. Schneider, Andrew W. Senior, John Jumper, Demis Hassabis, Pushmeet Kohli, and Žiga Avsec. Accurate proteome-wide missense variant effect prediction with AlphaMissense. Science, 381(6664):eadg7492, September 2023. ISSN 0036-8075, 1095-9203. doi: 10.1126/science.adg7492. URL https://www.science.org/doi/10.1126/science.adg7492.

